# Evaluation of different diffusion-weighted image techniques for head and neck radiation treatment: phantom and volunteer studies

**DOI:** 10.1101/2022.03.22.22272705

**Authors:** Yao Ding, Mohamed A. M. Meheissen, Kun Zhou, Abdallah S. R. Mohamed, Zhifei Wen, Sweet Ping Ng, Hesham M. Elhalawani, Baher A. Elgohari, Katherine A. Hutcheson, Caroline Chung, Stephen Y Lai, Clifton D. Fuller, Jihong Wang

## Abstract

**Purpose:** To quantitatively compare and assess the geometric distortion and apparent diffusion coefficient (ADC) accuracy of conventional single-shot EPI (SSEPI)-DWI, readout segmentation of long variable echo-trains (RESOLVE)-DWI, and BLADE-DWI techniques in both phantom and volunteer imaging studies.

**Methods:** Phantom measurements were obtained using the QIBA DWI phantom at 0° and at ambient temperature (~17°). DW images were acquired using SSEPI-DWI, RESOLVE-DWI, and BLADE-DWI. Image geometric distortion factors (compression and dilation, shear distortion), and image shift factors were measured and compared with computed tomography images. ADC and signal-to-noise ratio (SNR) values were measured for the various DWI techniques. Images were also obtained from three healthy volunteers and three oropharynx cancer patients. Geometric distortion parameters and ADC values were analyzed in the following head and neck regions of interest (ROI): salivary glands, tonsils, and primary tumors (patient only). The metric evaluation included Dice similarity coefficient and Hausdorff distance mean. T2-weighted images were used as a reference to evaluate geometric distortion.

**Results:** In the phantom experiment, the vials were prominently distorted on the SSEPI-DWI and RESOLVE-DWI images but were less distorted on the BLADE-DWI image, as determined by the overall effects of Image geometric distortion factors. The paired t-test showed no significant difference (0.15 < *p* < 0.88) in ADC values among the three DWI techniques; however, BLADE-DWI led to a lower SNR in vials. In the volunteer and patient studies, an ROI-based overlap metrics analysis of the salivary glands and gross tumor volumes were less distorted for BLADE-DWI than for EPI-based DWI, as determined by Wilcoxon paired signed-rank test.

**Conclusion:** BLADE-DWI demonstrated excellent geometric accuracy, with similar quantitative ADC values to those of EPI-based DW-MRI, thus potentially making this technique suitable for target volume delineation and functional assessment for head and neck radiation treatment in the future.

## 1. INTRODUCTION

It is crucial to accurately delineate the target (gross tumor) in radiation treatment planning. Particularly in head and neck cancer, computed tomography (CT)-based target delineation showed high interobserver variation ^1^whereas MRI-based target delineation tends to be more precise with much less interobserver variance. Diffusion-weighted magnetic resonance imaging (DW-MRI) is a functional imaging technique that provides information on the local microscopic mobility of water in tissue ^2^. Dense tissue with high cellularity, such as gross tumors and metastatic lymph nodes, restricts the motion of water and has high contrast in comparison to the surrounding normal tissue on high-b-value diffusion-weighted imaging (DWI) ^3^. For this reason, DWI has the great potential to be used to improve the accuracy of target delineation and assessing the treatment response for adaptive therapy.

Conventional DWI is based on single-shot echo planar imaging (SSEPI); however, this technique is prone to geometric distortion artifacts, especially in the head and neck region, because of significant magnetic susceptibility variation, which results from air-tissue or dental filling-tissue boundaries ^4^, thus, hindering the utility of SSEPI-DWI in target delineation.

An alternative to SSEPI-DWI is readout segmentation of long variable echo-trains (RESOLVE)-DWI. RESOLVE is a relatively new approach to DWI; it reduces sensitivity to local magnetic field inhomogeneity and may therefore lead to a substantial reduction in image distortion compared with the conventional SSEPI ^5^. However, this approach only partially eliminate image distortions, as the local magnetic field inhomogeneities around head and neck tumors are often strong.

BLADE-DWI is a multi-shot, turbo-spin echo-based DWI technique that is inherently insensitive to susceptibility-induced local magnetic field inhomogeneity ^6,7^. Inter-shot motion correction is applied by oversampling the region in the center of the k-space to reduce in-plane motion artifacts, which benefits image co-registration between different b-value acquisitions ^8 9^. Therefore, BLADE-DWI can substantially improve geometric accuracy and warrant the accuracy of ADC estimation.

Our hypothesis in this study was that the BLADE-DWI is superior to EPI-based DW-MRI techniques, including SSEPI DWI and RESOLVE DWI, in discriminating between target volumes (that is gross tumor) and normal structures in the head and neck region. Thus, we quantitatively compared the geometric distortion and ADC value accuracy of conventional SSEPI-DWI, RESOLVE-DWI, and BLADE-DWI techniques in head and neck imaging in both phantom and human subject studies.

## 2. METHODS AND MATERIALS

### 2.A. MRI Acquisition

MRI was performed on a MAGNETOM Aera 1.5T MR scanner (Siemens Healthcare, Erlangen, Germany) with two large four-channel flex phased-array coils.

#### 2.A.1. Phantom data acquisition and analysis

We first performed phantom studies to validate ADC accuracy and geometric distortion using the three DWI techniques studied: SSEPI-DWI, RESOLVE-DWI, and a work-in-progress (WIP) version of BLADE-DWI sequence developed by the vender. We also compared the signal-to-noise ratios (SNRs) of these three techniques. A QIBA DWI phantom (Model 128, High Precision Devices, Inc., Boulder, CO, USA) was used. It contains 13 vials filled with 30 ml of polymerpolyvinylpyrrolidone (PVP) in aqueous solution ^10^. These vials contained six different concentrations of PVP, corresponding to different ADC values. We measured the ADC values at 0°C (ice water bath) and 17°C (ambient temperature), respectively. The SSEPI-DWI scan was performed with QIBA-recommended parameters, whereas RESOLVE-DWI and BLADE-DWI protocols were individually optimized to match SSEPI-DWI image quality. The details of the acquisition parameters are presented in Table 1.

**Table 1:**
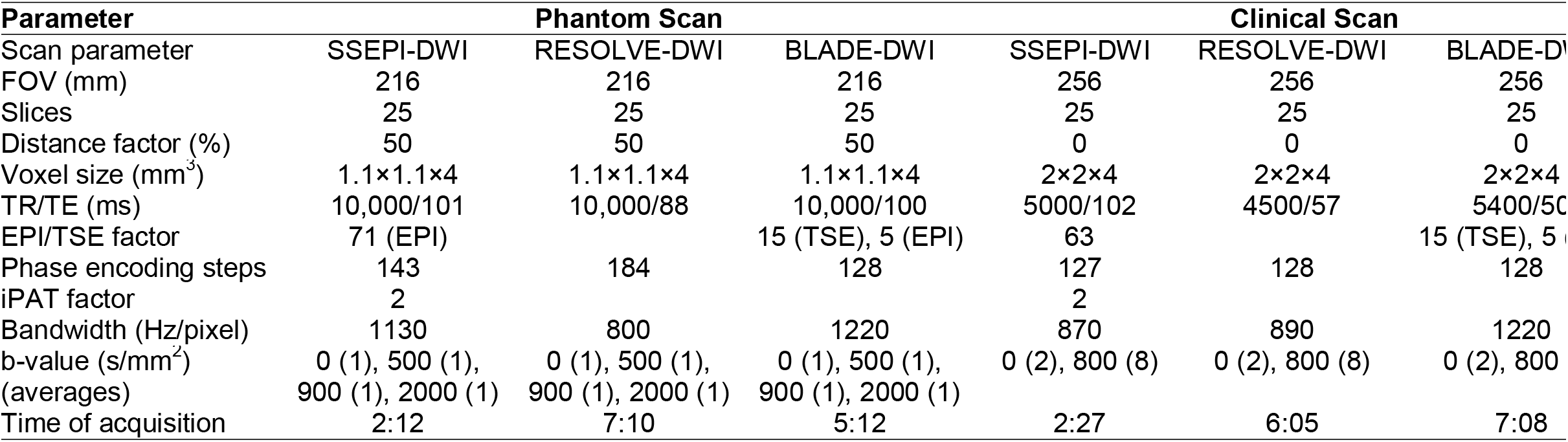
Sequence parameters (SSEPI-DWI, RESOLVE-DWI, and BLADE-DWI) used for the QIBA phantom and clinical imaging.

| Parameter | Phantom Scan |  |  | Clinical Scan |  |  |
| --- | --- | --- | --- | --- | --- | --- |
|  | SSEPI-DWI | RESOLVE-DWI | BLADE-DWI | SSEPI-DWI | RESOLVE-DWI | BLADE-DWI |
| Scan parameter |  |  |  |  |  |  |
| FOV (mm) | 216 | 216 | 216 | 256 | 256 | 256 |
| Slices | 25 | 25 | 25 | 25 | 25 | 25 |
| Distance factor (%) | 50 | 50 | 50 | 0 | 0 | 0 |
| Voxel size (mm<sup>3</sup>) | 1.1×1.1×4 | 1.1×1.1×4 | 1.1×1.1×4 | 2×2×4 | 2×2×4 | 2×2×4 |
| TR/TE (ms) | 10,000/101 | 10,000/88 | 10,000/100 | 5000/102 | 4500/57 | 5400/50 |
| EPI/TSE factor | 71 (EPI) |  | 15 (TSE), 5 (EPI) | 63 |  | 15 (TSE), 5 (EPI) |
| Phase encoding steps | 143 | 184 | 128 | 127 | 128 | 128 |
| iPAT factor | 2 |  |  | 2 |  |  |
| Bandwidth (Hz/pixel) | 1130 | 800 | 1220 | 870 | 890 | 1220 |
| b-value (s/mm<sup>2</sup>) | 0 (1), 500 (1), | 0 (1), 500 (1), | 0 (1), 500 (1), | 0 (2), 800 (8) | 0 (2), 800 (8) | 0 (2), 800 (8) |
| (averages) | 900 (1), 2000 (1) | 900 (1), 2000 (1) | 900 (1), 2000 (1) |  |  |  |
| Time of acquisition | 2:12 | 7:10 | 5:12 | 2:27 | 6:05 | 7:08 |

After obtaining all three diffusion weighted images, we calculated the ADC maps using a simple mono-exponential model on a pixel-by-pixel basis and in-house post processing software (Matlab, MathWorks, MA, USA). Regions of interest (ROIs) were contoured in the centers of vials on five middle slices; the means and standard deviations of ADC of each ROI were subsequently calculated for each technique (Fig. 1). The SNRs of the inner vial were estimated on the highest b-value (b = 2000 s/mm^2^) DWI. We compared diffusion images (b = 2000 s/mm^2^) with the corresponding reference CT images. We assessed geometric distortion using three factors—compression and dilation, shear distortion, and image shift—and ImageJ software (National Institutes of Health, Bethesda, MD, USA) ^11^. We then used the sum of three factors to evaluate overall geometric distortion (OGD) at the inner, middle, and outer layer vials (Fig. 2).

**Fig. 1.**
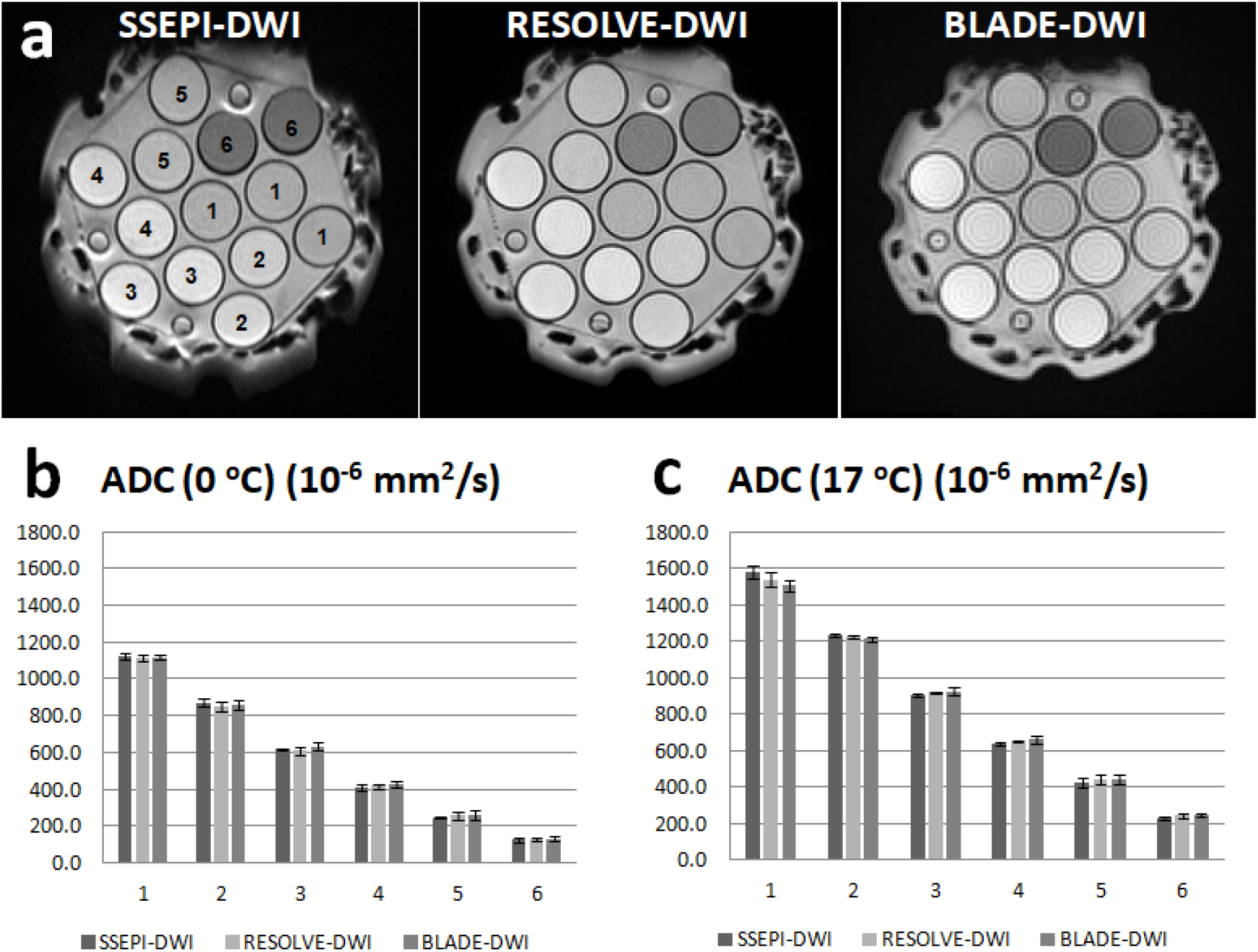
(a) DW images (b2000) of the ice QIBA phantom using different DWI techniques. The number indicates different concentrations of PVP in aqueous solution (the same number means the same concentration of PVP). (b, c) ADC values (with standard deviation) of SSEPI-DWI, RESOLVE-DWI, and BLADE-DWI at 0°C (b) and ambient temperature (17°C) (c).

**Fig. 2.**
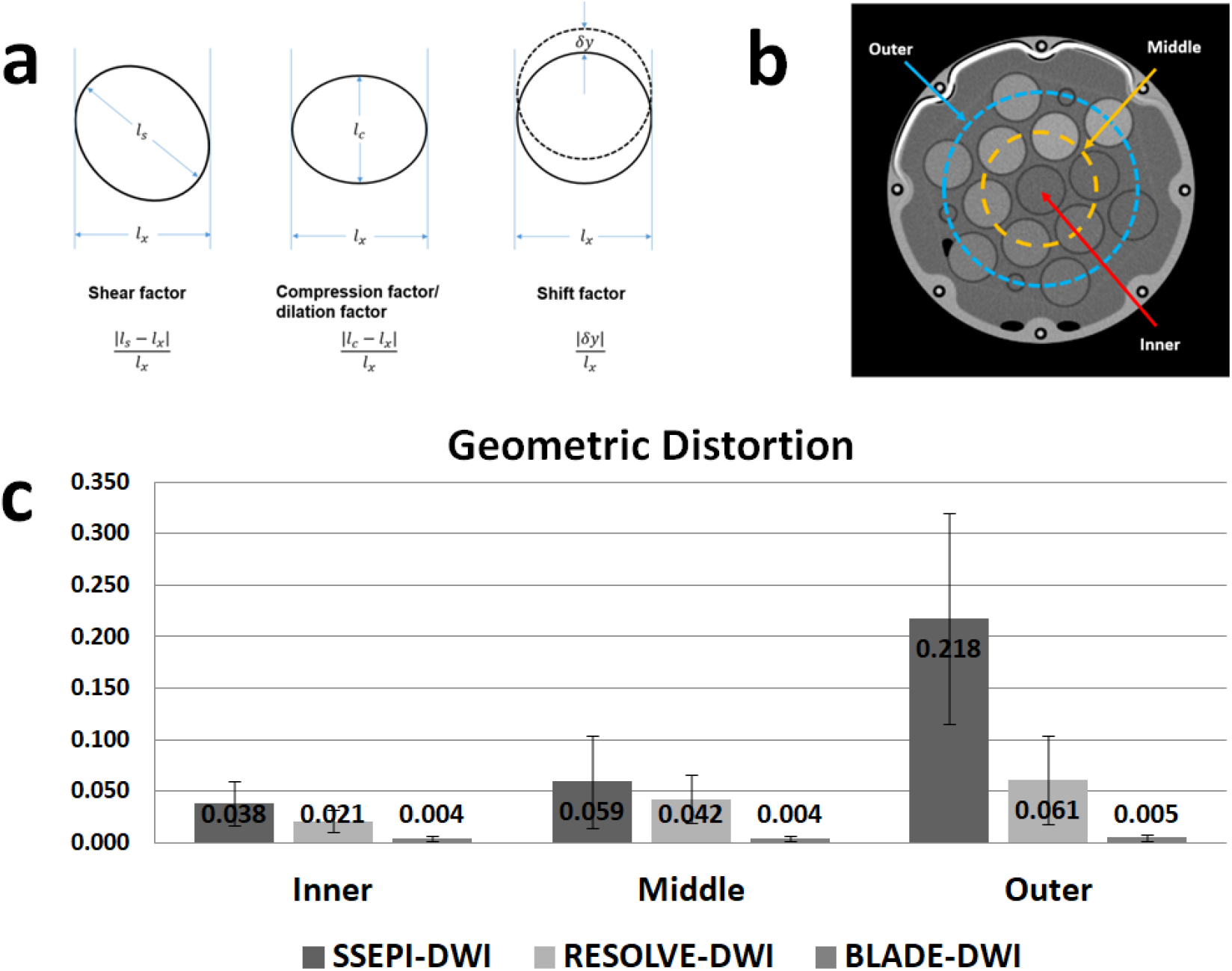
(a) A schematic diagram of the definition of geometric distortion factors: (1) shear, (2) compression or dilation, and (3) shift. (b) Layer definition shown on a CT image. (c) The overall geometric distortion factors (the sum of shear factor, compression factor, and shift factor) among SSEPI-DWI, RESOLVE-DWI, and BLADE-DWI.

#### 2.A.2. Volunteer and patient data acquisition and analysis

After receiving institutional review board approval for this prospective observational study, we enrolled three healthy volunteers and three patients with oropharyngeal cancer after obtaining signed study-specific informed consent forms to demonstrate the *in vivo* feasibility of using the BLADE-DWI technique to mitigate geometric distortion during head and neck MRI. Healthy volunteers underwent head and neck MRI in a thermoplastic mask that simulated the treatment position during radiation treatment. Patient MR simulation imaging was performed in the treatment position using the full set of immobilization devices, including a Klarity cushion head support (Klarity Medical Products, Newark, Oh, USA), thermoplastic mask, and mouth stent ^12^. We performed anatomical T2-weighted (T2W) imaging (FOV = 256 mm; matrix = 512 × 512; TR/TE = 6920/82 ms; ETL = 19; pixel bandwidth = 200 Hz; acquisition time = 6 min) and three DW-MRI techniques (SSEPI-DWI, RESOLVE-DWI, and BLADE-DWI) (Table 1).

All images were exported to Velocity software (Velocity AI 3.0.1; Velocity Medical Solutions, Atlanta, GA) for contouring and rigid-body registration. Contours of ROI were created by a senior radiation oncology resident and included the bilateral parotid and submandibular glands lingual tonsils in patients and volunteers and primary tumors in patients on T2W and DW images using the three techniques (b = 0 for normal gland delineation and b = 800 s/mm^2^ for gross tumor volume [GTV] delineation), independently. These contours were reviewed and approved by four radiation oncologists with 5-7 years of experience each.

Rigid registration was performed among the different DWI techniques and T2W images; ROIs contoured on individual sequences were propagated from the DW images to the T2W images. The propagated DWI volumes of interest were compared with those contoured on the anatomical T2W images using the Dice similarity coefficient (DSC) (conformality) and surface distance metrics (the Hausdorff distance mean [HDM]) for geometric distortion assessment. The geometric distortion (phantom and organ) of the three DWI techniques was compared using the Wilcoxon paired signed-rank test. Phantom ADC values among the three DWI sequences were compared using a paired t-test. *P* < 0.05 refers to a significant difference, while *p* < 0.01 indicates that the difference was extremely significant. All analyses were executed using JMP Pro version 11 software (SAS Institute, Cary, NC, USA).

## 3. RESULTS

### 3.A. QIBA phantom DWI evaluation

The geometric distortion of the QIBA phantom was more prominent in SSEPI-DW images than in RESOLVE-DW and BLADE-DW images. The layer vials from the inner layer to outer layer became prominently distorted on SSEPI-DW and RESOLVE-DW images but were consistently less distorted on BLADE-DW images (Fig. 2c). Specifically, the amount of geometric distortion on the middle and outer layers was substantially lower with BLADE-DWI (middle: OGD = 0.004; outer: OGD = 0.005) than with SSEPI-DWI (middle: OGD = 0.059; outer: OGD = 0.218) and RESOLVE-DWI (middle: OGD = 0.042; outer: OGD = 0.061). ADC values were measured for the three DWI techniques at six different PVP concentrations; we found no significant difference in these values (0.15 < *p* < 0.88), and the difference range among the three sequences was within 5% when ADC > 0.3 × 10^−3^ mm^2^/s (the reported ADC values of cancerous and noncancerous tissues were larger than 0.4 × 10^−3^ mm^2^/s) (Fig. 1) ^13,14^. However, the resulting SNR of the inner vial was lower with BLADE-DWI (SNR ~10) than with SSEPI-DWI (SNR ~30) and RESOLVE-DWI (SNR ~25).

### 3.B. Volunteer and patient DWI evaluation

DWI was successfully performed in all three healthy volunteers and three patients. The ROI-based overlap metrics analysis of the salivary glands, tonsils, and GTVs revealed significantly less distortion with BLADE-DWI (DSC:[0.72, 0.80]; HDM:[0.44, 0.83]) and RESOLVE-DWI (DSC:[0.65, 0.77]; HDM:[0.65, 1.17]) than with SSEPI-DWI (DSC:[0.41, 0.59]; HDM:[1.25, 2.43]) (Fig. 3). However, RESOLVE-DWI (DSC = 0.65; HDM = 0.81) was less geometrically accurate when used to image tonsils than was BLADE-DWI (DSC = 0.72; HDM = 1.17).

**Fig. 3.**
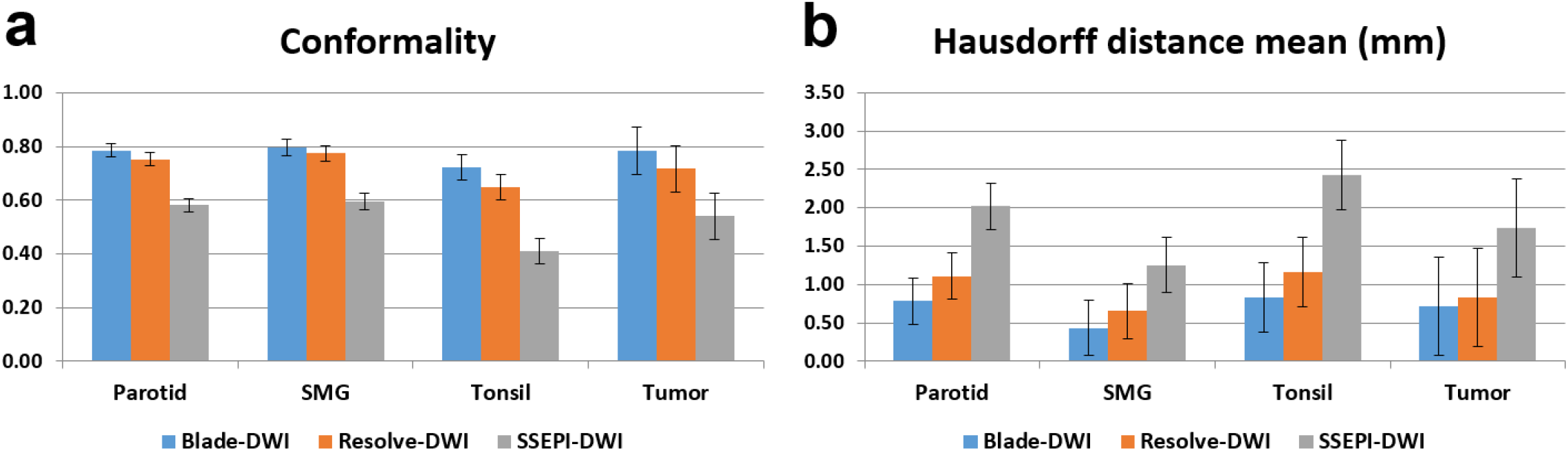
Assessment of geometric distortion factors. (a) Conformality and (b) the mean Hausdorff distance among BLADE-DWI, RESOLVE-DWI, and SSEPI-DWI.

An individual metrics comparison of the three sequences is detailed in Table 2. Fig. 4 shows DW images obtained using the three different techniques in a healthy volunteer. Image distortion is evident in the SSEPI-DWI scans (Fig. 4e) when the ROI delineation is compared with the reference T2W image delineation (Fig. 4b). Both the RESOLVE-DWI scan (Fig. 4d) and BLADE-DWI scan (Fig. 4c) demonstrated geometric accuracy on salivary glands (Fig. 4a). However, compared to BLADE-DWI, RESOLVE-DWI demonstrated a higher degree of geometric distortion, particularly in structures close to air cavities (tonsils). Fig. 5 shows an example of GTV delineation among the three diffusion techniques. The image distortions around the air cavities are easily recognized in the SSEPI-DWI and RESOLVE-DWI scans (Fig. 5a, 5d, and 5e), while the BLADE-DWI scan (Fig. 5a and 5c) shows improved GTV accuracy. Diffusion contrast was present in all three DWI scans, with the tumor appearing hyperintense on b800 images.

**Table 2:** Organ geometric distortion comparisons for DWI techniques using Wilcoxon methods.

| Organ | Conformality (p-value) |  |  | Hausdorff distance mean (p-value) |  |  |
| --- | --- | --- | --- | --- | --- | --- |
|  | SSEPI vs.<br>BLADE | SSEPI vs.<br>RESOLVE | BLADE vs.<br>RESOLVE | SSEPI vs.<br>BLADE | SSEPI vs.<br>RESOLVE | BLADE vs.<br>RESOLVE |
| Parotid (12) | **0.012** | **0.015** | 0.653 | **0.026** | **0.024** | 0.762 |
| SMG (11) | **0.021** | **0.038** | 0.753 | **0.023** | **0.031** | 0.842 |
| Tonsils or tumor (12) | **0.007** | **0.009** | 0.243 | **0.008** | **0.011** | 0.461 |

SMG: Submandibular gland.

**Fig. 4.**
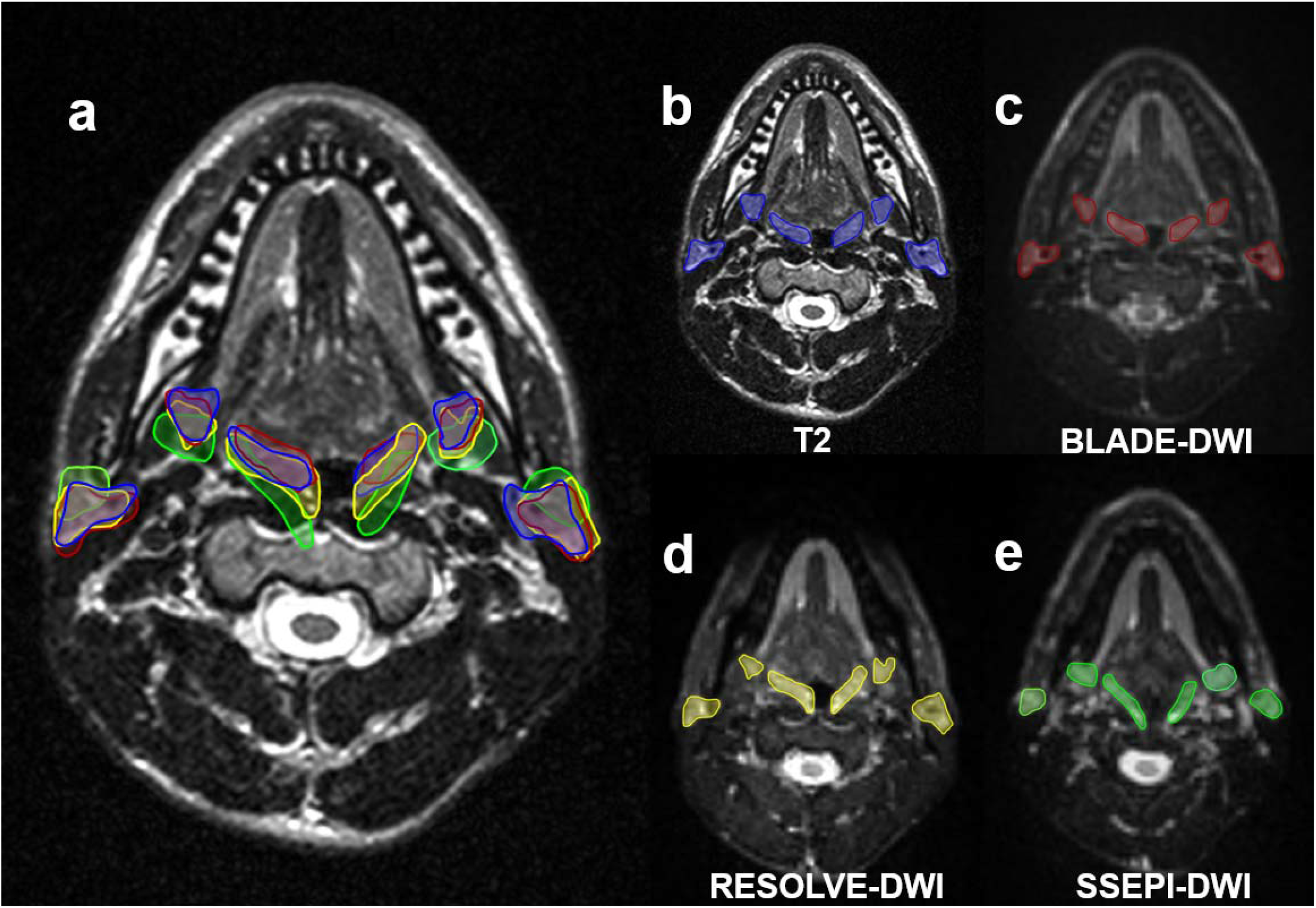
Example of DWI in a healthy volunteer. (a) T2W image with fused organ contours, including the submandibular glands, parotid glands, and tonsils. These delineated organs are indicated by a blue line on (b) the T2W image, a red line on (c) the BLADE b0 image, a yellow line on (d) the RESOLVE b0 image, and a green line on (e) the SSEPI b0 image.

**Fig. 5.**
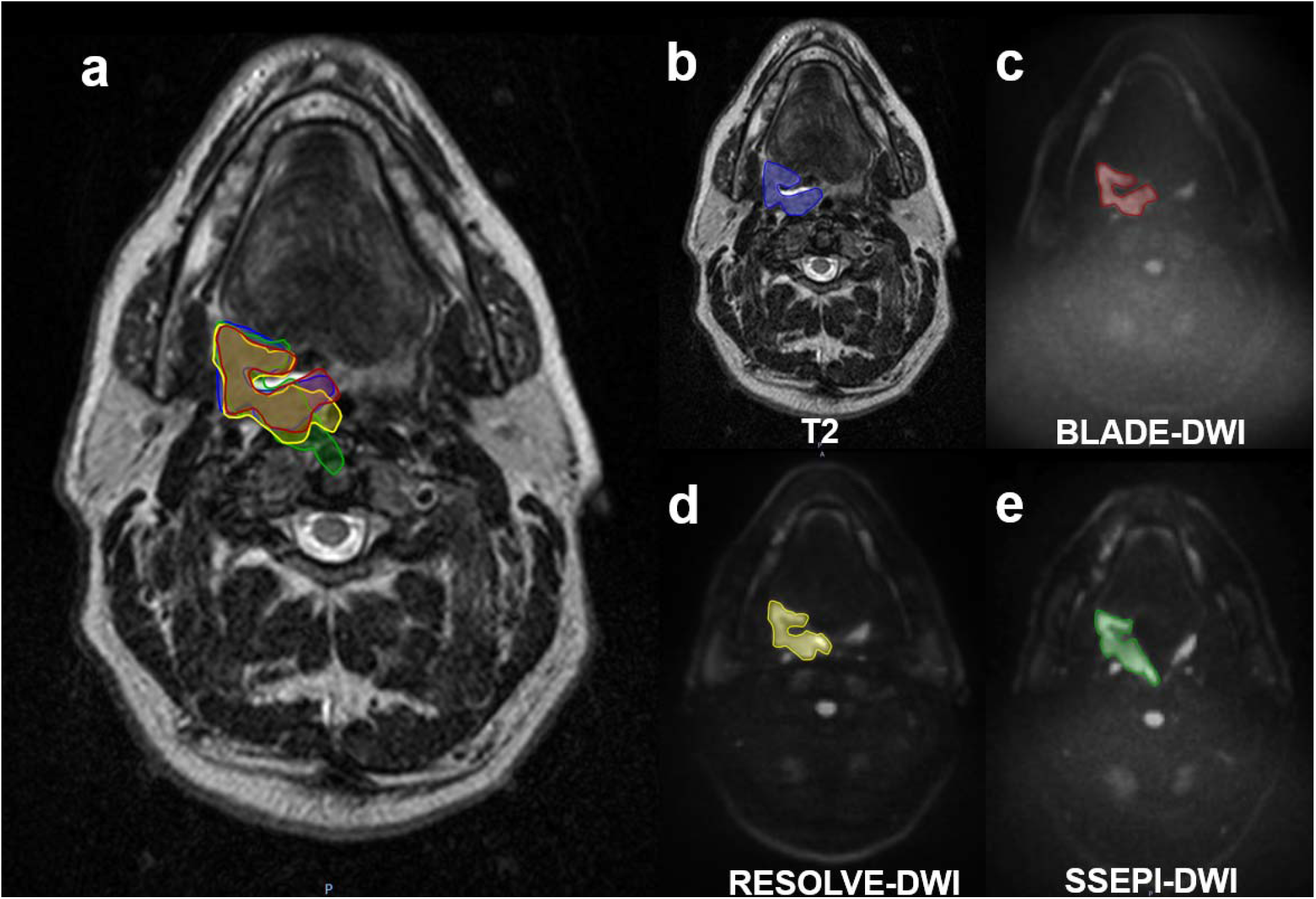
DWI in a representative patient. (a) T2W image with fused GTV overlap. The delineated GTV is indicated by a blue line on (b) the T2W image, a red line on (c) the BLADE b800 image, a yellow line on (d) the RESOLVE b800 image, and a green line on (e) the SSEPI b800 image.

## 4. DISCUSSION

DWI is a well-established functional MRI technique that have been used mainly for diagnosing cancer. It has not been incorporated extensively into radiation treatment planning because DW images obtained using SSEPI-based techniques are susceptible to large geometric distortion, especially in the head and neck region, as a result of complex anatomy and large variance of tissue susceptibility (bone, air cavity, and soft tissue). Therefore, a distortion-free DWI technique is needed to precisely delineate critical structures and lesions during radiation treatment planning. Furthermore, the ADC parameter has been shown to be a good clinical biomarker for assessing the effects of radiation treatment’ ^15,16^. Thus, it is necessary to verify the ADC accuracy of different DWI approaches.

In this study, we evaluated a BLADE-DWI technique at 1.5 T and compared it with EPI-based DWI techniques (SSEPI-DWI and RESOLVE-DWI); the BLADE-DWI sequence offered marked improvements in image quality, primarily by suppressing geometric distortions in the head and neck region. In addition, there was no significant difference in ADC values among the different DWI techniques. Therefore, BLADE-DWI appears to be an advantageous approach to head and neck radiation treatment planning and assessment. In the phantom study, our results also showed that BLADE-DWI was significantly less distorted, especially in the outer layer, which is under large susceptibility difference conditions. In the human subject studies, the findings were similar in ROIs in the head and neck region, where severe susceptibility artifacts are expected (table 2., tonsils and base of tongue tumors).

As far as we are aware, this is the first study to quantitatively compare the geometric accuracy of BLADE-DWI, SSEPI-DWI, and RESOLVE-DWI using phantom and clinical data. The difference in distortion between BLADE-DWI and SSEPI-DWI was striking on almost all ROI contours. While the geometric accuracy of BLADE-DWI was similar to that of RESOLVE-DWI for the parotid and submandibular glands, significantly higher image distortion was seen in the tonsils and base of tongue tumors on RESOLVE-DWI. This is likely because the susceptibility of parotid and submandibular glands is similar to that of the surrounding soft tissue, which will cause less distortion on RESOLVE-DW images. Structures located close to the air cavities may be prone to poorer geometrical accuracy. This superiority of BLADE-DWI over EPI-based DWI (SSEPI-DWI and RESOLVE-DWI) is crucial in multiple radiation treatment applications, including pre-treatment GTV definition, adaptive treatment planning, and post-treatment discrimination of residual tumor from radiation-induced normal tissue toxicity in head and neck squamous cell carcinoma.

One of the difficulties associated with BLADE-DWI, aside from its longer acquisition time, is the reduction in the SNR, which affects ADC value estimation ^17^. In our comparison of BLADE-DWI and EPI-based DWI sequences in the QIBA phantom, we observed a lower SNR with BLADE-DWI than with SSEPI-DWI and RESOLVE-DWI, when matched parameters were used. Therefore, in the volunteer study, we optimized the DWI sequences with a clinically acceptable time (Table 1, ~7 minutes for BLADE-DWI, ~6 minutes for RESOLVE-DWI, and 3 minutes for SSEPI-DWI) to ensure sufficient SNR for tumor target delineation and ADC calculation. An additional concern with the BLADE technique is the high specific absorption rate (SAR) because of the use of multiple refocusing pulses in a fast-spin echo mode. This SAR limitation may confine the number of slices that can be fit within a given TR, which might limit the application of BLADE-DWI in some cases of large coverage imaging applications (e.g., from the skull base to low neck region). In this study, a vendor-supplied WIP package was used, which made it possible to use a multi-blade k-space filling strategy. This efficient multi-echo acquisition technique allowed us to reduce the specific absorption rate deposition and accelerate data acquisition.

This study has some limitations. The study included only three patients with gross tumors; therefore limiting the power to detect significant differences in the ADC values between BLADE-DWI and EPI-based DWI. However, the QIBA phantom study in 0°C and ambient temperature showed no significant difference of ADC values between these techniques. Further studies with larger numbers of patients are required to validate the ADC accuracy in head and neck cancers. We did not test the short-term repeatability and reproducibility of BLADE-DWI, so as not to inconvenience the patients. However, efforts are under way to perform daily or weekly BLADE-DWI to test our measurements in other protocols.

## Conclusions

BLADE-DWI was more geometrically accurate than were SSEPI-DWI and RESOLVE-DWI in both the phantom and human subject studies, without significant difference of ADC values. Thus, BLADE-DWI technique can be potentially used to facilitate functional assessment, with accurate target delineation, in head and neck cancer radiation treatment planning and assessment which might benefit radiomic analysis.

## Data Availability

All data produced in the present study are available upon reasonable request to the authors.

## Acknowledgements

The authors thank the Proton Treatment Center MRI team (Shane Ikner and Vi Dinh) for their helps in acquisition of MR studies. We would like to thank Ann Sutton at the Department of Scientific Publications of MD Anderson Cancer Center for her editorial support in the preparation of this manuscript.

